# Development and Validation of the Cannabis Exposure in Pregnancy Tool (CEPT)

**DOI:** 10.1101/2022.09.09.22279777

**Authors:** Kathleen H. Chaput, Carly A. McMorris, Amy Metcalfe, Catherine Ringham, Stephen Wood, Deborah McNeil, Shaelen Konschuh, Laura Sycuro, Sheila W. McDonald

**Affiliations:** Department of Obstetrics and Gynecology, Cumming School of Medicine, University of Calgary; Department of Community Health Sciences, Cumming School of Medicine University of Calgary; Werklund School of Education, School and Child Psychology, University of Calgary; Maternal Newborn Child and youth Strategic Clinical Network, Alberta Health Services; Research and Innovation Population, Public, and Indigenous Health, Alberta Health Services; Department of Microbiology, Immunology and Infectious Diseases, Cumming School of Medicine, University of Calgary

## Abstract

**Background:** Evidence of associations between prenatal cannabis use (PCU) and maternal and infant health outcomes remains conflicting amid broad legalization of cannabis across Canada and 40 American states. A critical limitation of existing evidence lies in the non-standardized and crude measurement of PCU, resulting in high risk of misclassification bias. We developed a standardized tool to comprehensively measure prenatal cannabis use in pregnant populations for research purposes.

**Methods:** We conducted a patient-oriented tool development and validation study using a bias-minimizing process. Following an environmental scan and critical appraisal of existing prenatal substance use tools, we recruited pregnant participants via targeted social media advertising and obstetric clinics in Alberta, Canada. We conducted individual in-depth interviews and cognitive interviewing in separate sub-samples, to develop and refine our tool. We assessed convergent and discriminant validity internal consistency and 3-month test-retest reliability, and validated the tool externally against urine THC bioassay.

**Results:** 254 pregnant women participated. The 9-item Cannabis Exposure in Pregnancy Tool (CEPT) had excellent discriminant (Cohen’s kappa=-0.27-0.15) and convergent (Cohen’s kappa=0.72-1.0) validity; as well as high internal consistency (Chronbach’s alpha = 0.92), and very good test-retest reliability (weighted Kappa=0.92, 95% C.I. [0.86-0.97]). The CEPT is valid against urine THC bioassay (sensitivity=100%, specificity=77%).

**Interpretation:** The CEPT is a novel, valid and reliable measure of frequency, timing, dose, and mode of PCU, in a contemporary sample of pregnant women. Using CEPT (compared to non-standardized tools) can improve measurement accuracy, and thus the quality of PCU and maternal and child health research.

## Background

Admist a backdrop of cannabis legalization, prenatal cannabis use (PCU) is rising (1,2). Despite recent studies showing associations between PCU and adverse maternal, infant, and child outcomes (3–6), the evidence remains conflicting (7–12). A critical limitation of published studies is a high risk of misclassification bias resulting from a lack of standardized measurement of PCU across adequate domains, including frequency, dose, modes, timing of use in pregnancy, and second-hand smoke and vapour. There is an urgent need for high-quality cannabis-related health research, and pregnant individuals and infants have been identified as priority populations (9,10,13). Improved measurement of PCU in research is a key component to improving the quality of the evidence.

Current PCU measurement options available for research include administrative data collected during routine prenatal care, substance use disorder (SUD) screening tools, non-validated questionnaires, and biological tests. Administrative data is problematic for research use because pregnant people are known to under-report prenatal substance use to physicians (14,15). Further, PCU screening is not standardized practice, occurs variably, and is seen as low-priority for the majority of obstetricians (16). While Canadian studies using administrative data have reported PCU prevalence between 2% and 3% (2–4), emerging evidence from an *anonymous* population-based survey indicates an 11% prevalence of PCU (17). In a US study only 36% of women with cannabis-positive urine tests had reported their use to a care provider (18), indicating that the majority of prenatal cannabis users may be misclassified in administrative data studies.

While self-administered research questionnaires can garner more accurate reporting of substance (e.g. alcohol) use in pregnancy than screening in clinical settings (19,20), non-standardized survey questions have limited utility for measurement of PCU, as they can unintentionally convey perceived bias against PCU. They often identify cannabis as an illicit drug and do not differentiate between medicinal and recreational use, which contradicts social perception and may increase response bias (18,21). Survey questions are problematic for studying nuanced associations with maternal and infant health outcomes due to inconsistent assessment of frequency and timing of use, including changing patterns through pregnancy, and often lack dose measurement, or use subjective dose-terminology (9,10,22–28). Further, most lack measurement of potentially important consumption modes aside from smoking (vapourized, edible, topical, second-hand)(22–24). Standardized SUD screening tools aim to detect a diagnosable SUD, and do not measure patterns PCU throughout pregnancy (29). Many screen for alcohol misuse alone (30–33), or combine all drugs into a single category (29) preventing the separate evaluation of cannabis-related health outcomes. Biological (urine/blood/saliva) cannabis-screeners exist, but are limited to detection within 1-5 weeks of use, depending on individual metabolism and test cut-off levels (34–38). Biological samples are also resource-intensive and stigmatizing to collect, limiting their utility for prospective research. Our study developed and validated a novel PCU measurement tool, that addresses the limitations of current measurement methods, using a patient-oriented approach to identify patient-perceived stigma, and reduce perceived sources of response bias, using a six-step, peer-reviewed process (39).

## Methods

We recruited pregnant, past- and current-cannabis users and non-users, between 08/2019 and 04/2020 for the mixed-methods tool development phase and an external validation cohort between 04/2022 and 12/2022. We used social media advertising targeted to women aged 18-45 years, residing in Alberta, with listed interests or group memberships related to pregnancy, parenting, and/or cannabis, and posted gender-neutral recruitment ads in an online transgender parent support group. Study recruitment letters were also mailed to patients who visited Alberta Health Services (AHS) clinics for pregnancy-related care in the preceding six months, identified using pregnancy-related codes in the National Ambulatory Care Reporting System (NACRS)(Appendix A). We included participants meeting target criteria who were <36 weeks’ gestation at intake. Our target development sample size of 150 participants was sufficient to detect a Cronbach’s alpha of ≥0.9, with 95% confidence for test-retest reliability on a tool that contains up to 15 items (39), and our external convenience sample of 85 participants was feasible for conducting urine THC bioassays with available resources. This study was approved by the Conjoint Health Research Ethics Board at the University of Calgary (REB19-0670), and written informed consent was obtained from all participants.

### Step 1 Qualitative Interviews

We conducted individual in-depth interviews with 10 regular and occasional cannabis users, and non-users, purposively selected from the full sample (Figure 1). Two research assistants with qualitative interview training conducted telephone interviews at a time chosen by the participant, about views and experiences with cannabis use in general, and during pregnancy. Prior to interviews, research staff contacted participants twice to discuss study details, including confidentiality, and establish a trusting relationship, by disclosing their own connections to the study topic, emphasizing a non-judgmental approach, and acknowledging all experiences shared were important. We recorded and transcribed interviews verbatim, and used deductive thematic analysis to extract pre-determined themes of: language around cannabis and its use; perceptions of stigma and judgement, and their relationships to truthful disclosure of use; patterns of use in pregnancy (timing, frequency of use, typical dose); motivations for use; and forms of cannabis used. Two team members experienced in qualitative methods coded salient content that corresponded to the pre-determined themes, collapsed codes into broader themes using constant comparison technique, discussion and consensus. Themes were then reported back to the qualitative participants via email for member-checking of the relevance and appropriateness to ensure truth value.

**Figure 1:**
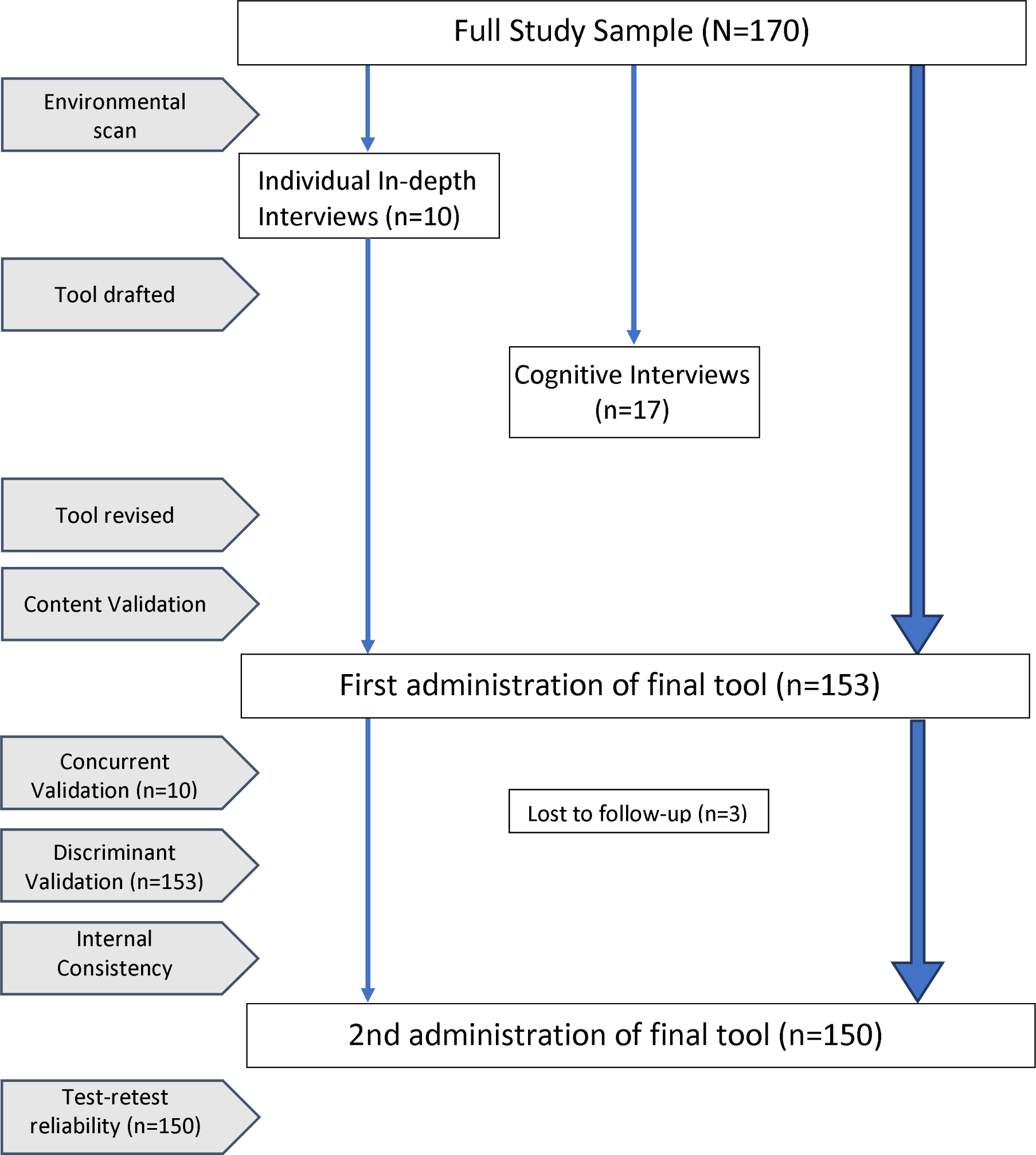
Study Flow Diagram: Development phase

### Step 2 Devising Items

We devised items, including wording, to draft the tool based on strengths and shortcomings identified in existing SUD tools and published survey questions (Table 1), and on themes identified from interviews. We eliminated double-barreled questions, ambiguous wording and ensured a 6th grade reading level.

**Table 1:** Measurement domains of existing prenatal cannabis measurement options.

| *Includes measure of:* | **4ps** | **4ps+** | **WIDUS** | **CRAFFT** | **SURP-P** | **StatsCan** | **Generation-R** | **NSDUH** |
| --- | --- | --- | --- | --- | --- | --- | --- | --- |
| *Cannabis separately* | no | no | no | yes | yes | yes | yes | yes |
| *Use in pregnancy* | no | yes | no | No | no | no | yes | *indirect |
| *Frequency of use* | no | no | no | no | no | yes | yes | yes |
| *Timing of use in pregnancy* | no | no | no | no | no | no | 1<sup>st</sup> trimester & pre-preg. | *indirect |
| *Dose* | no | no | no | no | no | yes | no | no |
| *Mode of consumption* | no | no | no | no | no | yes | no | no |
| *Medicinal vs. recreational use* | no | no | no | no | no | yes | no | no |
| *Time-span covered* | Past ever | Past month | Past ever | Past 12 months | Past ever | Past 3 months | Pre-pregnancy, First trimester | Past 3 mos. |
| *Second-hand exposure/partner use* | yes | yes | yes | no | yes | no | yes | no |
| *Reference* | (29) | (30) | (31) | (32) | (33) | (44) | (5) | (50) |

*4Ps=Parent drug problem, Partner drug problem, Past use of substance*
*4Ps+= Parent drug problem, Partner drug problem, Past use of substance, Pregnancy use*
*WIDUS= Wayne Indirect Drug Use screener; CRAFFT = Car, Relax, Alone, Forget, Friends, Trouble;*
*SURPP= substance use risk in pregnancy profile; NSDUH = National Survey on Drug Use and*
*Health; StatsCan = Statistics Canada \* specific date can be cross-referenced with pregnancy*
*information if provided*

### Step 3 Cognitive interviewing and bias reduction

Schwartz and Oyserman (40) propose five stages of cognition required to accurately self-report behaviour, each of which are susceptible to bias: 1. question understanding, 2. recalling relevant behaviour, 3. inference & estimation, 4. mapping answer onto response options, and 5. answer editing. To identify points of bias at all five stages of cognition, we conducted individual *cognitive interviews* with an additional sub-sample of participants from the full sample, in which respondents were asked to think aloud, and share impressions, understanding, and reasoning related to each of the five stages of cognition, as we administered the newly developed tool (41). We iteratively revised items according to participant feedback prior to each subsequent interview, until no new suggestions for revision were made in two consecutive interviews (after interview 17).

### Step 4 Content Validation

We then formatted the refined items into the CEPT online tool, compared to our critical apporaisal of existing tools to ensure it captured all domains of measurement that are critical to prospective research cannabis in pregnancy, including timing, multiple modes of consumption, dose per use and frequency of use.

### Step 5 Convergent and Discriminant Validation

We then administered the finalized CEPT, along with the SURP-P(42) and 4Ps+ (30) SUD screening tools via electronic questionnaire, to our remaining sample of 150 women. We measured concurrent validity of CEPT responses against detailed cannabis use information revealed during the interviews using Cohen’s weighted kappa. There is strong evidence that a high degree of truth value can be achieved with rigorous qualitative interview techniques.(43) We assessed discriminant validity of CEPT responses against SURP-P and 4Ps+ tools using Cohen’s kappa. We calculated internal consistency on all CEPT cannabis consumption items using Chronbach’s alpha, acknowledging that it measures multiple constructs of cannabis exposure (i.e. any use, frequency, timing, dose, mode and reasons), rather than a single construct. However, we anticipated internal consistency among the CEPT items, as a person indicating use should have non-zero responses for dose, mode frequency and reasons for use. We then re-administered the tool to all development-phase participants (n=150) 3 months later to assess test-retest reliability using a weighted Cohen’s kappa (Figure 1).

### Step 6 External validation

In an additional external sample of 84 pregnant participants, we validated CEPT responses against urine bioassay measurements of 11-nor-9-carboxy-Δ^9^-THC, the most abundant THC metabolite (Figure 2). Participants provided urine samples in sterile collection containers that were shipped frozen to our laboratory by pre-paid courier for analysis, within 24 hours of completing an online questionnaire including the CEPT. We stored samples at −80°c until analysis. 2ml aliquots were taken from thawed samples, centrifuged and diluted (10x) with ultrapure water and assayed in duplicate using a 96-strip-well, THC Metabolite ELISA Kit (catalogue # 701570, Cayman Chemicals^TM^, United States of America) according to manufacturer’s protocol. No freeze-thaw cycles were allowed, and the lowest threshold of THC positivity detectable by the kits (0.072*η*g/ml) was used to classify those with PCU versus those without.

**Figure 2:**
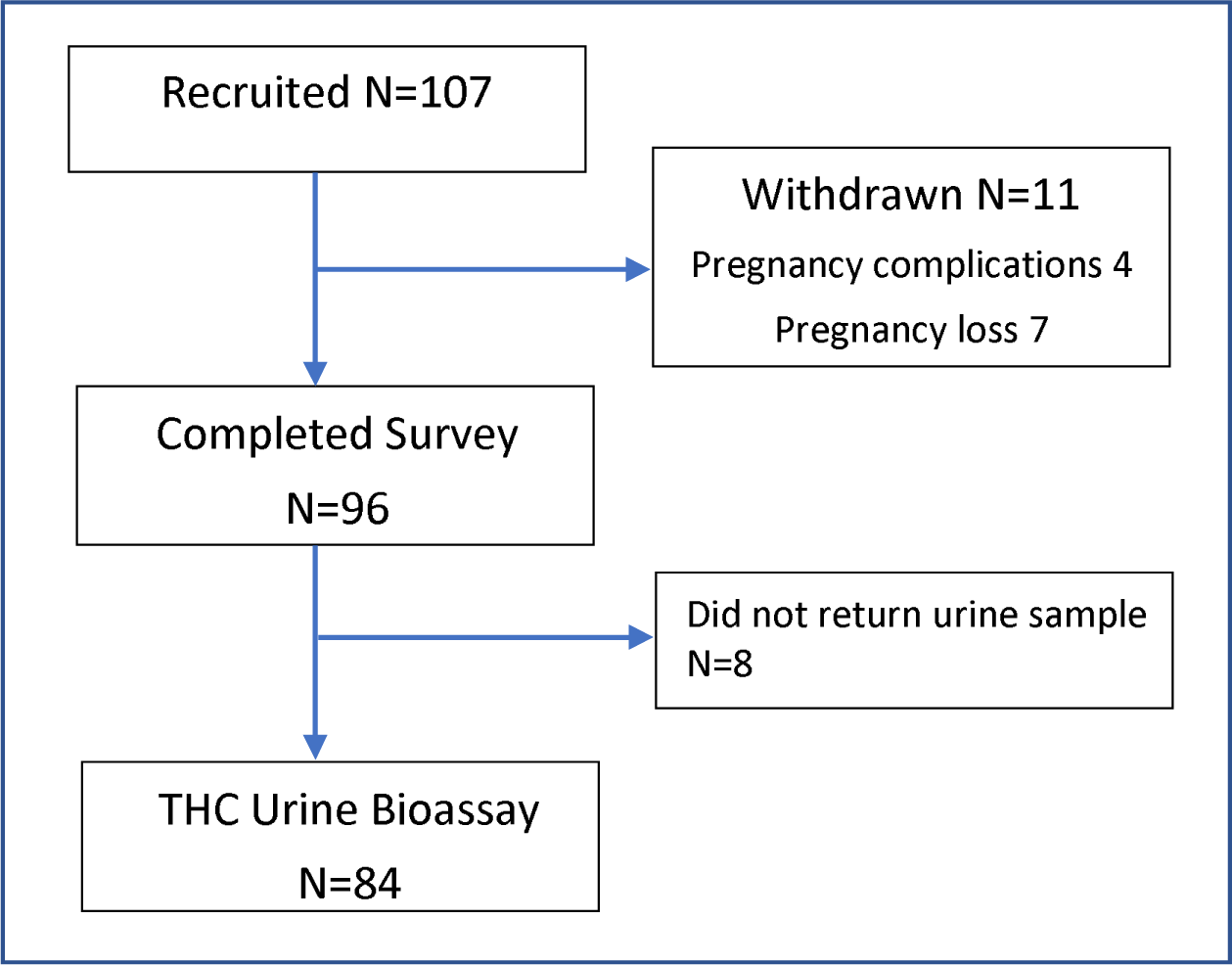
Study Flow Diagram – External Validation.

## Results

Our sample included 254 pregnant past, current, and non-consumers of cannabis, 170 in the development phase and 84 in the external validation cohort. Specific sub-samples participated in various steps (Figures 1,2). Table 2 summarizes participant characteristics at enrollment. Other sociodemographic characteristics of our sample were similar to the overall maternal population in Canada (44–46). (Fig. 3)

**Figure 3:**
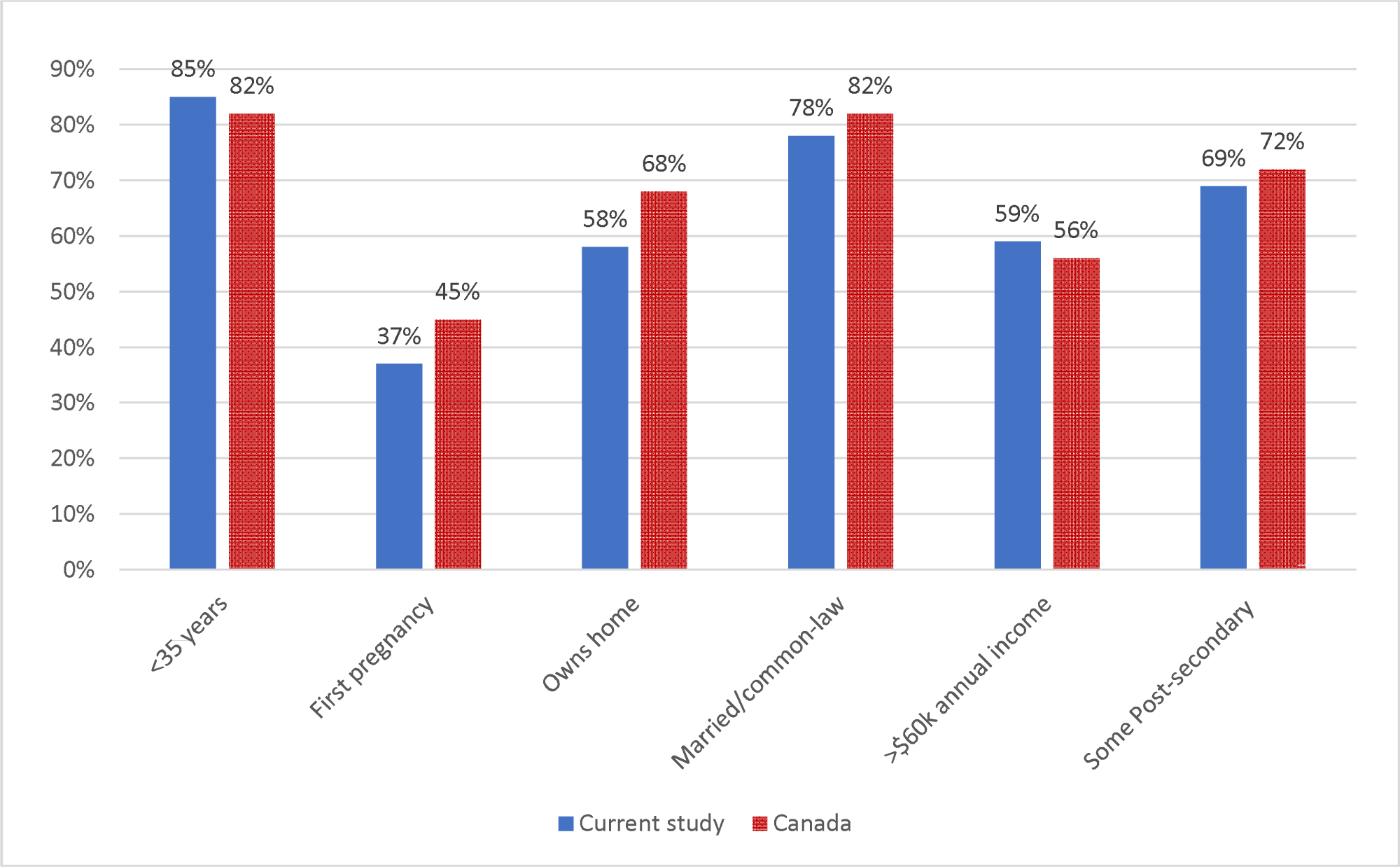
Participant Characteristics Versus Maternal Population of Canada.

**Table 2:**
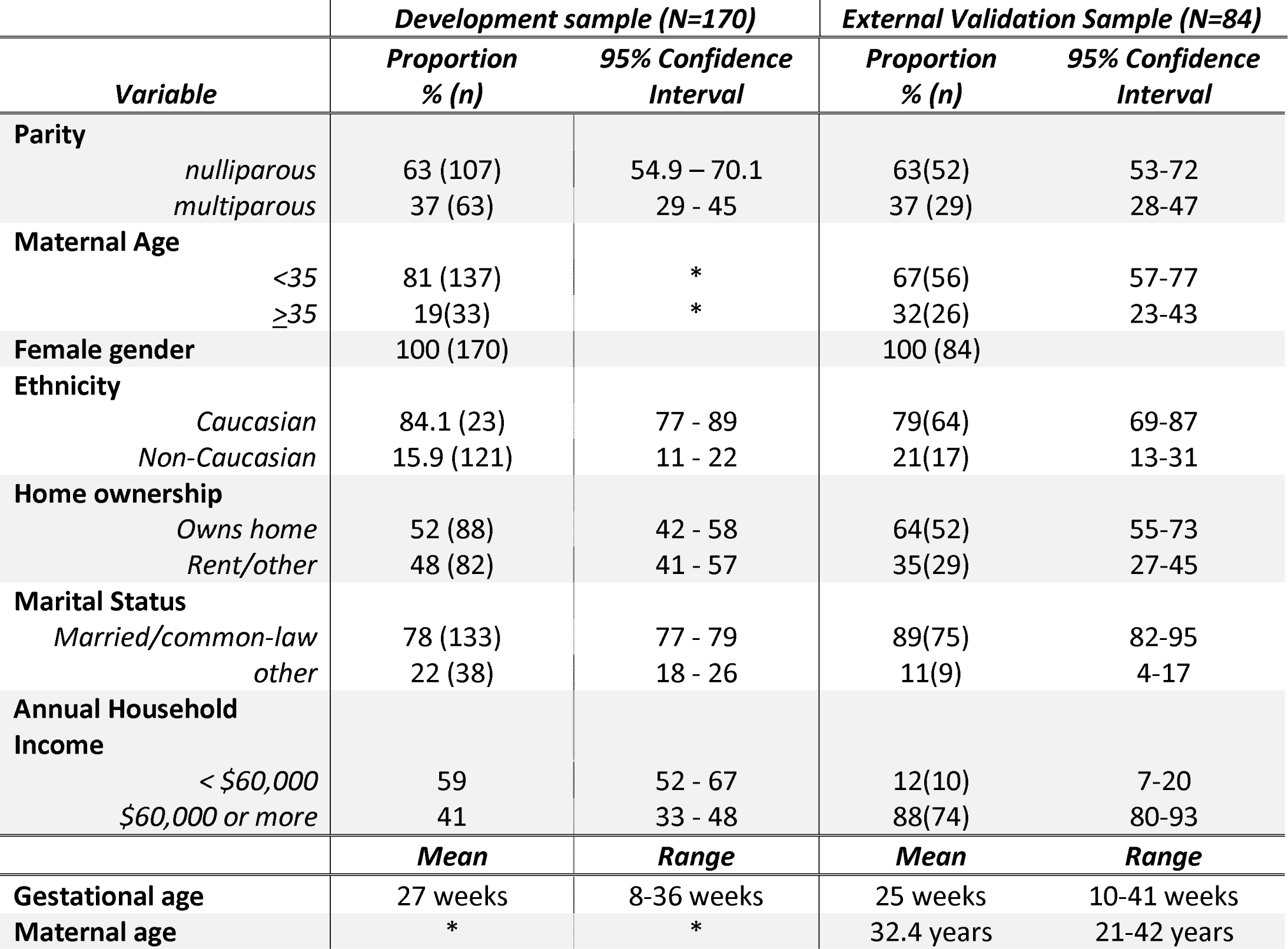
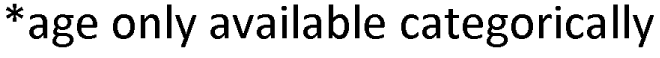
Participant Characteristics at Enrollment.

| **Variable** | **Development sample (N=170)** |  | **External Validation Sample (N=84)** |  |
| --- | --- | --- | --- | --- |
|  | **Proportion**<br>**% (n)** | **95% Confidence**<br>**Interval** | **Proportion**<br>**% (n)** | **95% Confidence**<br>**Interval** |
| **Parity** |  |  |  |  |
| nulliparous | 63 (107) | 54.9 – 70.1 | 63(52) | 53-72 |
| multiparous | 37 (63) | 29 - 45 | 37 (29) | 28-47 |
| **Maternal Age** |  |  |  |  |
| <35 | 81 (137) | * | 67(56) | 57-77 |
| ≥35 | 19(33) | * | 32(26) | 23-43 |
| **Female gender** | 100 (170) |  | 100 (84) |  |
| **Ethnicity** |  |  |  |  |
| Caucasian | 84.1 (23) | 77 - 89 | 79(64) | 69-87 |
| Non-Caucasian | 15.9 (121) | 11 - 22 | 21(17) | 13-31 |
| **Home ownership** |  |  |  |  |
| Owns home | 52 (88) | 42 - 58 | 64(52) | 55-73 |
| Rent/other | 48 (82) | 41 - 57 | 35(29) | 27-45 |
| **Marital Status** |  |  |  |  |
| Married/common-law | 78 (133) | 77 - 79 | 89(75) | 82-95 |
| other | 22 (38) | 18 - 26 | 11(9) | 4-17 |
| **Annual Household**<br>**Income** |  |  |  |  |
| < \$60,000 | 59 | 52 - 67 | 12(10) | 7-20 |
| \$60,000 or more | 41 | 33 - 48 | 88(74) | 80-93 |
|  | **Mean** | **Range** | **Mean** | **Range** |
| **Gestational age** | 27 weeks | 8-36 weeks | 25 weeks | 10-41 weeks |
| **Maternal age** | * | * | 32.4 years | 21-42 years |

\*age only available categorically

### Qualitative interviews

We completed qualitative data collection after 10 interviews, when we reached thematic saturation (no new themes emerged). Summaries of deductive themes and illustrative quotes are presented in Table 3. Interviews informed bias-minimizing language and wording, tool structure, and appropriate response options for frequency dose and reasons for use. Themes drove the terminology and language used in the tool preamble and questions, guided tool structuring including inclusion of specific items (e.g. reasons for use) and response options, and determined the method of dose measurement. While legalization was perceived to have reduced stigma around cannabis use in general, perceptions of stigma against prenatal use were prevalent and thus important for consideration to encourage accurate disclosure. Several participants noted that including a response option to disclose cannabis consumption that occurred only prior to pregnancy recognition was crucial, and noted if this option was not present, they would not report use, even if they had consumed cannabis prior to pregnancy recognition. A challenging aspect of cannabis consumption measurement is identifying dose. IDI results identified a reliable method of quantifying approximate dose per use (i.e. comparing amounts to common objects, like food items or coins). Approximate THC/CBD content can be inferred based on mean THC content of dried cannabis available on the contemporary market (24%)(47), or the labeled concentration of products as reported by participants.(Supplementary file 2)

**Table 3:** Deductive themes and illustrative quotes *(n=10)*

|  |  |
| --- | --- |
| **Theme 1 - Language/ wording:** Participants indicated that non-judgemental wording around cannabis use as well as specific terms and context affected their choice to disclose their cannabis use while pregnant. |  |
|  | “...say “cannabis” instead of marijuana, because I think of marijuana only being the plant... not edibles, or cbd and lip balm, or whatever.” |
|  | “Why you want to know is important. I would be way more up-front if I know it’s for research, than like, if they want to know at the hospital...” |
|  | “If someone sounds judgmental, that would make me not want to discuss it. If it’s something that it’s clear that they’re open to it, ... I would be like, yeah, here’s how I take it and why.” |
|  | “I wouldn’t say ‘use’, I would say ‘consume’. [use] has a bit of a negative to it.” |
|  | “[marijuana] sometimes has a negative connotation, like it’s a drug, but cannabis is more ... like it’s natural.” |
| **Theme 2a) Tool structure - General:** The need for non-judgemental wording, and for cannabis to be treated separately from other substances on a questionnaire were identified as essential to avoid biased responses. |  |
|  | "Say something at the beginning to make it clear that you're not judging. If it sounds judgmental, or like, if I think you're asking me so you can lecture me ... someone's judging me for using it, I wouldn't answer." |
|  | "I feel like if doctors were a little non-judgmental and a little less biased, then it would create some more honesty." |
|  | "If it's lumped in with, you know, smoking, then drinking, then marijuana, then ... heroin and cocaine, that just gives it a real negative tone... like, it's worse than alcohol, and almost as bad as heroin... I wouldn't be answering, really, if it's like that." |
|  | "it makes more sense to me to have it with... supplements, or alternative therapies." |
| **Theme 2b) Tool structure - Response options:** Allowing participants to indicate their reasons for consuming cannabis in pregnancy (which were predominantly reported as medicinal), was perceived as a key factor for encouraging honest disclosure. A response option indicating that use only took place prior to pregnancy recognition was also seen as essential to unbiased reporting. |  |
|  | "I believe the stigma has died a lot. But there is still a big stigma with pregnancy for some people." |
|  | "Especially in the first pregnancy women feel a lot more judged." |
|  | "I don't believe it should be used in pregnancy to get stoned, or to get high. But I believe that if it's going to help with morning sickness, or relieve pain, or anything that you're going through that may cause you suffering or stress, I believe it safe to use..." |
|  | "some people stop as soon as they find out [they're pregnant], so you need to be able to say that." |
| **Theme 3a) Patterns of use – Mode of consumption:** Participants indicated numerous modes of consumption (vapour, oral/edible, topical, cannabidiol (CBD)) with varying doses for each, and some perceived as safer in pregnancy than others, supporting the need for standardized measurement of consumption-routes beyond smoking. |  |
|  | "I would think that ingesting it... would be a lot safer [than smoking] because there's less transfer to the fetus." |
|  | "I think edibles and lotions and liquid CBD capsules even, they're most likely more safe to take during pregnancy considering just that you're taking out the smoking out of the equation" |
|  | "I don't necessarily think that smoking it is the smartest." |
|  | "I mostly smoke, but I have drops and a lotion too." |
|  | "... for vaping it, [I] stick to three puffs maximum when it comes to THC products." |
|  | "Smoking does work quite quickly, especially for morning sickness. But a tincture can work..." |
|  | "I consume CBD oil daily, as well as smoking [cannabis]." |
| **Theme 3b) Patterns of use – Frequency and Timing:** Participants consistently indicated their patterns of use changed during pregnancy to a more frequent consumption of smaller amounts, compared to their general use pre-pregnancy, indicating that tool response options need to include high frequencies (i.e. multiple times per day) and small dose-per-use categories, compared to existing survey questions. |  |
|  | "I use it different (sic) now that I'm pregnant... I have a quick drag whenever I need it, so 3 or 4 times a day sometimes, but just a tiny bit, instead of having a lot at once." |
|  | "... I resumed micro-dosing daily..." |
|  | "I think asking about frequency makes sense - most people use it pretty regular (sic)" |
| **Theme 3b) Patterns of use – Dose:** Amounts of cannabis typically consumed at each sitting was discussed primarily in subjective terms (i.e. large, small), perceptions of which may vary considerably between consumers, and identifying the weights or exact doses used at each sitting was perceived as difficult or infeasible, particularly for dried cannabis. Comparison measures were preferred. |  |
|  | "I know how much I buy by weight, but I couldn't tell you the grams I put in the pipe yesterday evening..." |
|  | "With smoking it, it's harder... like, a big joint for me might not be big for my sister." |
|  | "It's easy if it's an edible, because it tells you on the label..." |
|  | "The THC oil that I have is 30 milligrams per mil, so that would work out to being about point three milligrams for point one or point two of a milliliter." |
|  | "Maybe start at half a milliliter, so that would be what? More like 10 milligrams, I guess, of the 40 milligram [per milliliter] stuff that I have." |
|  | "I would say the easiest way for people to say how much they smoke would be like a pea-sized amount, or a grape-size...compare it to something. Then you could figure out the grams from that. I don't know how many grams or milligrams I use every time." |
|  | "I might use a small dab like the size of a dime, or other times it might be like twice as much..." |

### Cognitive interviews

We completed cognitive interviews with a separate sub-sample of 17 participants to assess and minimize points of bias through participant-led refinement (Figure 4). This resulted in 9 sequential iterations of our initial draft tool. Perceived sources of bias at all five stages of cognition were identified, and changes made based on participant feedback.

**Figure 4:**
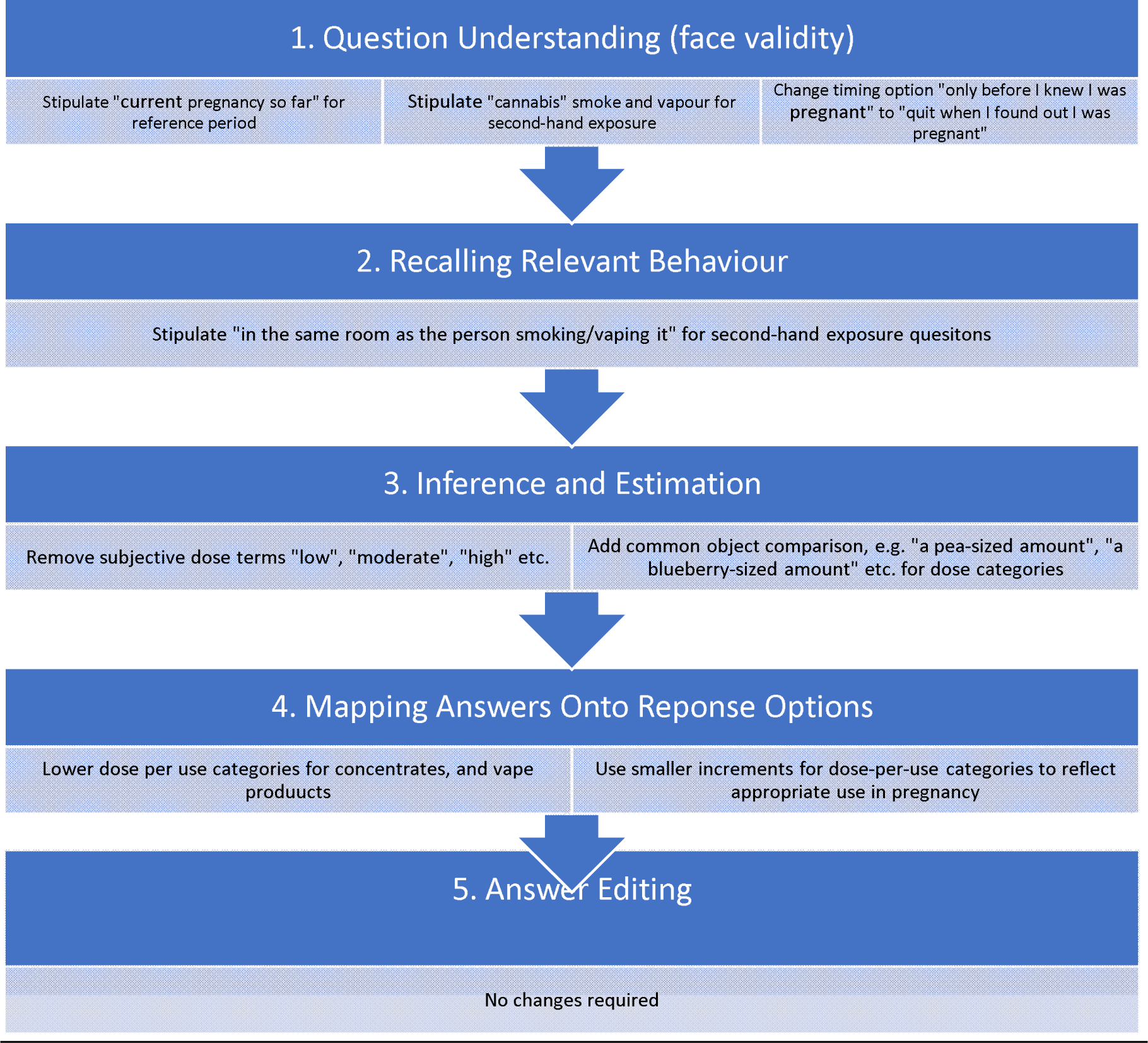
Cognitive Interviews - Bias reduction for the five stages of cognition.

#### Question understanding

Most draft-tool questions were well understood; however, some changes were made to improve clarity.

#### Recalling relevant behaviour

All participants indicated they were accurately able to recall details of first-hand cannabis consumption, including frequency, trimester of consumption, reasons, modes, and amounts per use. Nearly all participants (93%) indicated they were able to accurately recall the details of second-hand cannabis smoke or vapour exposure, aside from brief outdoor exposures. We amended the second-hand exposure question to include exposure while in the same room as the user.

#### Inference & estimation

Participants did not express concerns about inference or estimation on items measuring any consumption/exposure, or frequency, timing or reasons for use. Dose questions were adjusted to address perceived ambiguity and aid with estimation (Figure 4). *Mapping answers onto response options:* Several participants noted problems with initial dose-per-use options, increments for some product types were deemed too large for use in pregnancy, and we refined categories to align with appropriate ranges and increments. *Answer editing:* No participants expressed the need to edit responses once the above clarifications and response-option edits had been made. Participants agreed the tool was non-judgemental, appropriate, and acceptable to them in pregnancy, and that it would elicit truthful responses, confirming face and content validity from the participant perspective.

The final CEPT has 9 items measuring weeks of gestation, second-hand exposure, partner use, trimester(s) of use, frequency, reasons, modes of consumption, and dose per use for each mode indicated. Frequency, reasons, modes, and dose items repeat for each trimester of use indicated. (Appendix A)

### Validity and reliability

Concurrent validity was excellent, with agreement between IDI participant CEPT responses and use reported in IDIs, ranging from 80% to 100%, and kappa values ranging from substantial (0.72) to perfect (1.0) (49) (Table 4). The timing of use construct showed the lowest level of agreement, which was expected. Use will be reported in more trimesters as a pregnancy progresses. A greater proportion of participants (40%) reported third-trimester use on the online CEPT, compared with IDIs (30%), which occurred 5-6 weeks prior. Discriminant validation indicated poor agreement between two pregnancy SUD screening tools (5ps and the SURP-p)(33), with weighted Kappa values ranging from −0.31 to 0.36 indicating that the CEPT measures different constructs from those on the existing tools. (Table 5)

**Table 4:** Concurrent validity of the CEPT vs. In-depth interview (*n=10)*

| **Construct** | **Agreement** |  | **Kappa** | **Std. Error** | **P-Value** |
| --- | --- | --- | --- | --- | --- |
|  | *actual* | *expected* |  |  |  |
| *Any use in pregnancy* | 100% | 82% | 1.00 | 0.31 | >0.001 |
| *Frequency of use* | 90% | 22% | 0.87 | 0.16 | >0.001 |
| *Timing (trimester)* | 80% | 28% | 0.72 | 0.19 | >0.001 |
| *Mode of consumption* | 100% | 22% | 1.00 | 0.21 | >0.001 |

**Table 5:**
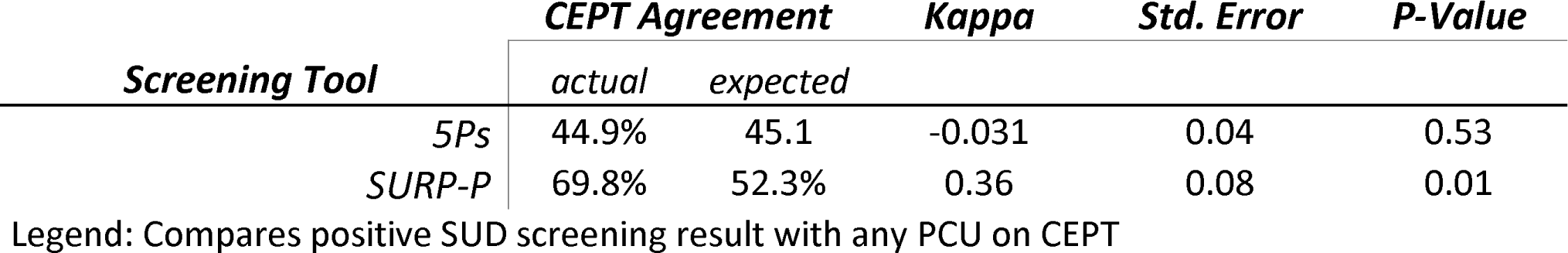
Discriminant validity of CEPT versus SUD screening tools (n=153)

Reliability testing showed excellent internal consistency (Chronbach’s alpha=0.92) and substantial to near-perfect Kappa values (0.71-0.99) for test-retest reliability (Table 6). Although some patterns of use may be expected to change throughout pregnancy, the strong agreement between early and late pregnancy responses on the CEPT support that recall of cannabis consumption using this tool is reliable up to delivery.

**Table 6:** Test-retest reliability of the CEPT - 3-month interval (n=153)

| **Construct** | **Agreement** |  | **Kappa** | **Std. Error** | **P-Value** |
| --- | --- | --- | --- | --- | --- |
|  | *actual* | *expected* |  |  |  |
| *Any second-hand exposure* | 92% | 52% | 0.83 | 0.08 | >0.001 |
| *Any use in pregnancy* | 97% | 51% | 0.95 | 0.08 | >0.001 |
| *Frequency of use* | 90% | 28% | 0.86 | 0.04 | >0.001 |
| *Timing (trimester)* | 80% | 32% | 0.71 | 0.05 | >0.001 |
| *Mode of consumption* | 99% | 51% | 0.97 | 0.08 | >0.001 |

CEPT-reported cannabis use was valid against urine-THC bioassay with 100% sensitivity, and 77% specificity, indicating that it has promise as an improved measure of PCU for research purposes (Table 7). All participants with positive urine bioassay disclosed cannabis use within 1 week of the urine sample being collected, indicating that the time elapsed since last use was the main driver of lower specificity.

**Table 7:**
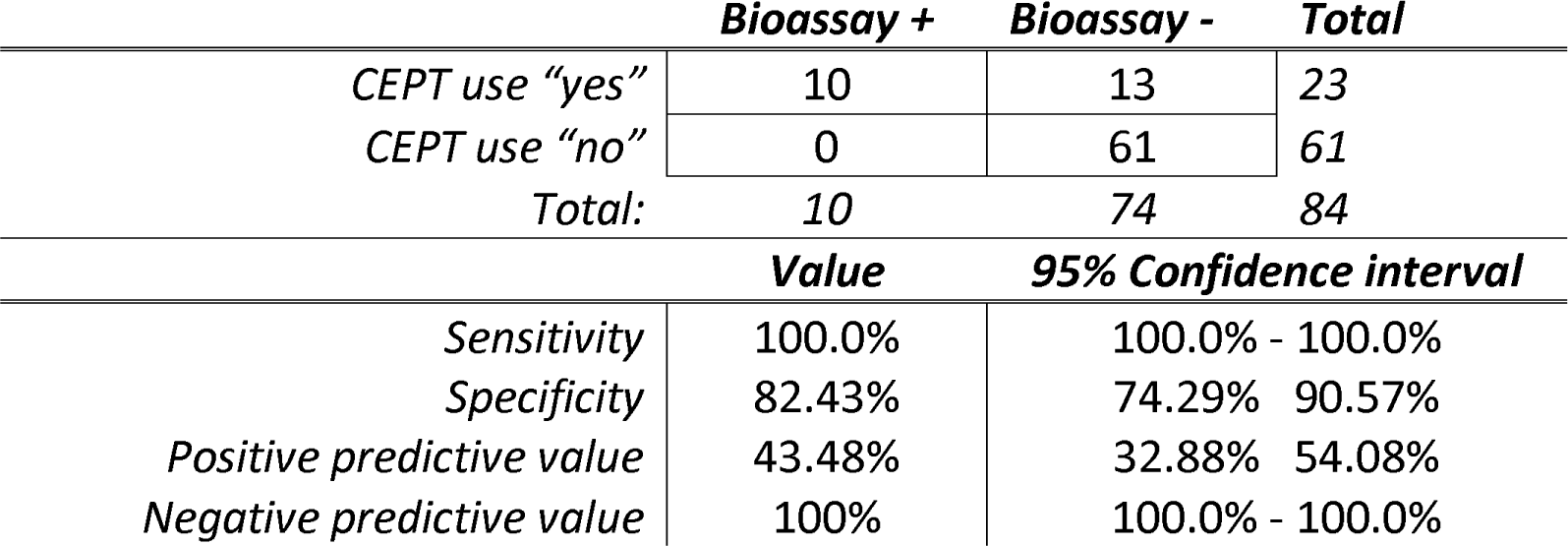
External Validation.

### Interpretation

The CEPT addresses the measurement limitations faced by previously published studies of PCU and maternal and infant health, which are highly susceptible to misclassification bias, have inconsistent findings, and are rated moderate at best by the US National Academies of Science Engineering and Medicine(10,50). It offers researchers a measurement option with strong validity and reliability, that accounts for frequency, modes, reasons and estimated dose-per-use, separately measures CBD and THC, and allows repeated measures per trimester to capture changing patterns of PCU. It alos measures frequency and timing of second-hand exposure, in addition to partner’s cannabis use. The CEPT thus enables more complete picture of exposure over pregnancy than currently published studies have been able to capture. The patient-oriented methods we used are a strength; qualitative interviews can reveal aspects of health behaviour that contrast with the researcher’s underlying assumptions, that can interfere with the five stages of cognition leading to biased response (39,40). Prenatal alcohol use studies indicate that non-disclosure bias for prenatal substance use varies according to participant perceptions, and that question wording and structure informed by patient-oriented designs can improve validity (20,51). Further, the language, tone, and perceived intent of the tool are critical to non-biased response. Our qualitative interviews guided us in reducing perceived judgemental or stigmatizing language in our tool. The cognitive interviews further reduced sources of bias. While we may never be able to completely eliminate PCU reporting bias our patient-oriented development process was chosen because it is crucial for minimising perceived stigma, and ensuring a much lower probability of bias than the methods of measurement used in previous studies, including self-selection for biological samples, which do not allow the participant to explain their reasons for use, nor to judge the researachers’ motivations. Although there remains no feasible gold-standard measure of prenatal cannabis consumption across the entire gestational period, the CEPT represents a useful tool for researchers to augment the quality and expand the scope of longitudinal research into the health outcomes associated with prenatal cannabis exposure. Our results support that it minimizes self-report bias, and its nuanced measurement of multiple dimensions of cannabis consumption may also reduce misclassification of very low exposures, allow for assessment of potential dose-response relationships, and enable the identification of critical windows of fetal exposure in future studies, that were not possible with previous crude measures.

#### Limitations

The CEPT is designed to measure behaviours over pregnancy, rather than to detect a condition or health state. Where medical screening tools can be validated against diagnostic tests or interview, validating a measure of behaviour is more complex. A limitation of our study is the lack of a true gold-standard measure of PCU for validation, which was financially infeasible for this study, as it requires multiple bioassays of at least weekly serial urine samples throughout gestation. However, we have preliminarily validated CEPT responses against a biological reference-standard, showing excellent sensitivity and high specificity. While we could not attain a true biological gold-standard in our study, the validation we conducted against single bioassays, and in-depth qualitative interviews remains rigorous. Biological levels of THC metabolite cannot be falsified, and the qualitative methods we employed result in high credibility and truth-value for qualitative results (57). Further, interviews allowed for comparison of binary cannabis use as well as PCU patterns (modes, frequency, timing) that cannot be validated with a biological test. Although our study sample was adequate to detect a Cronbach’s alpha of ≥0.9 on a tool with up to 15 items, we acknowledge that our external bioassay validation sample (n=84) was small. Future validation studies should include larger samples to confirm findings. It is also important to note that our tool and the validation conducted are limited to English-speaking individuals, and translations will require further validation.

## Conclusion

PCU and its associated health outcomes have been identified as priorities for research in Canada and the U.S. following cannabis legalization (9). We recommend the CEPT as a rigorous, feasible, patient-oriented health research tool for measuring PCU. The use of the CEPT as a standardized measure of PCU in future studies can contribute substantial new knowledge about the implications of timing, dose, frequency, and modes of exposure for maternal, fetal, infant and child health, accounting for varying patterns of consumption and the strength and diversity of cannabis products available on the contemporary legal market. The CEPT has the potential to significantly improve measurement accuracy and thus the quality of research in this area, which can in turn inform evidence-based education, prevention and health policy to mitigate potential health risks.

Data sharing: Quantitative data can be made available in accordance with the ethics approval for the study, on reasonable request to the corresponding author.

## Data Availability

All quantitative data produced in the present work will be available upon reasonable request to the authors, within ethical constraints upon completion of analyses. Qualitative data cannot be released for ethical reasons.

